# Risk Assessment of Unregulated Biological Sample Handling Among Postgraduate Students: A Mixed-Methods Study

**DOI:** 10.1101/2025.09.20.25336242

**Authors:** Abdulwahid M. Dhahir, Sara A. Nagmesh

## Abstract

This mixed-methods study assessed the risks of unregulated biological sample handling among postgraduate students in Iraq. Quantitative data, gathered from a single survey completed by 172 participants and analyzed descriptively, revealed widespread challenges in obtaining samples (71.5%) and accessing equipped laboratories (78.5%), driving students to rely on commercial labs (65.1%) and informal peer exchange (51.7%). Twenty-five semi-structured interviews further elucidated these findings, revealing underlying reasons such as material shortages and poor university lab infrastructure (mentioned in 21 interviews). Both methodologies indicated high risk awareness (91.3% of survey respondents acknowledged risks; 16 of 25 interviewees recognized them), yet students continue these practices out of necessity. Interviews particularly emphasized a severe lack of adequate and practical biosafety training (indicated by 15 interviews). Crucially, in the absence of specialized governmental biosecurity agencies in Iraq, these unregulated practices heighten the risk of sensitive biological materials reaching unauthorized hands or being misused.

## Introduction

Biological samples, such as tissues, blood, and bodily fluids, serve as the cornerstone of scientific advancement and innovation across medicine, pharmacology, and life sciences. The safe and regulated handling of these samples is paramount not only for ensuring the integrity of scientific research and the accuracy of its findings, but also for safeguarding the health and safety of individuals who handle them, and the wider community (Lakin & Capell, 2014) (World Health Organization, 2004). Any lapse in biological sample management practices poses a potential risk, ranging from contamination and data loss to direct health hazards associated with exposure to pathogenic agents (Ragavan et al., 2023; Sveinbjornsson & Gizurarson, 2022a). The concepts of Biosafety and Biosecurity are gaining increasing prominence within academic and research environments, particularly in universities conducting extensive research involving sensitive biological materials (Morse, 2025a). Biosafety encompasses the principles, technologies, and practices implemented to prevent unintentional exposure to pathogens or toxins, or their release into the environment (Ragavan et al., 2023). Conversely, Biosecurity focuses on protecting valuable or dangerous biological materials from theft, diversion, misuse, or intentional release (Artika & Ma’roef, 2018). Unregulated practices, or those lacking adequate oversight, directly threaten both concepts, creating an environment where samples, personnel, and the community are put at risk.

In many research settings, especially in developing countries or those facing infrastructural challenges, postgraduate students, who constitute a vital part of the research workforce, may encounter significant limitations in accessing adequate laboratory resources and appropriate training (Badr, 2018). These challenges can compel them to adopt informal or unregulated practices in handling biological samples, such as relying on commercial laboratories, exchanging samples with colleagues, or even working in inadequately equipped environments (Verstuyft et al., 2018). While such practices may be driven by the necessity to complete research, they open the door to serious risks concerning sample integrity loss, contamination, mismanagement, and potential exposure to infectious agents (Chauhan & Jindal, 2020).

Iraq, as an academic and research environment with unique challenges, serves as a prominent example of a context where such practices can emerge. Following decades of conflict and limitations on investment in research infrastructure (Dakhil et al., 2024), Iraqi academic institutions may be in urgent need of a comprehensive assessment of biological sample handling practices, particularly among postgraduate students who often operate under significant time and financial pressure (Alrasheed et al., 2024a). The current scientific literature notably lacks comprehensive studies exploring this phenomenon specifically within the Iraqi context.

Building on the foregoing, this study aims to bridge this knowledge gap by comprehensively assessing the risks associated with the unregulated handling of biological samples among postgraduate students in Iraq. By employing a mixed-methods design that integrates both quantitative and qualitative data, this research seeks to provide an in-depth understanding of the prevalence of these practices, the underlying reasons, students’ risk perceptions, their level of biosafety awareness, and the overall impact on biosecurity. The anticipated findings will underscore the urgent need for developing effective policies and training programs to enhance biosafety and biosecurity practices within Iraqi academic institutions.

## 3. Methodology

### 3.1 Study Design

This research employed a mixed-methods design to provide a comprehensive understanding of the unregulated handling of biological samples among postgraduate students in Iraq. The study combined descriptive quantitative analysis, conducted through an online questionnaire, with qualitative analysis, derived from semi-structured interviews.

### 3.2 Data Collection Tools

Data were collected using both quantitative and qualitative instruments, as follows:

#### 3.2.1 Quantitative Data Collection

An online questionnaire was developed to gather quantitative data. It comprised four main sections: (1) Demographics, (2) Sample handling practices, (3) Risk assessment, and (4) Training and awareness. To ensure the reliability of responses, a trap question was included to detect inattentive or random answers. The instrument’s content relevance was further ensured through validation by subject matter experts.

#### 3.2.2 Qualitative Data Collection

Semi-structured interviews were conducted to gather in-depth qualitative insights. This approach balances a pre-determined set of topics or open-ended questions with the flexibility to explore emerging themes and follow up on participants’ responses in depth. Participants were selected using a quasi-purposive sampling method, focusing on postgraduate students with relevant experience or knowledge in biological sample handling practices. To maintain participant comfort and privacy, interviews were not audio-recorded. Instead, the researcher manually documented responses during the interviews and immediately reviewed them afterward to ensure accuracy and completeness.

### 3.3 Sampling Strategy

A non-probability sampling approach was utilized for both components of the study:

#### Quantitative Component

A convenience sample of postgraduate students was recruited from various medical and life sciences fields across several Iraqi universities. Proportional representation was not a requirement for this component.

#### Qualitative Component

Participants for the interviews were selected using a quasi-purposive sampling method, based on their direct relevance to the research topic and their willingness to share their experiences.

### 3.4 Data Analysis

Different analytical approaches were applied to the quantitative and qualitative datasets:

#### Quantitative Data Analysis

Quantitative data were analyzed solely using descriptive statistics, specifically frequencies and percentages. Inferential statistical tests were not applied due to the imbalanced representation of universities within the sample, which could introduce bias into the conclusions.

#### Qualitative Data Analysis

Qualitative data, derived from the interview responses, were analyzed using thematic analysis. This approach facilitated the identification of recurring themes and key insights pertinent to the research objectives.

### 3.5 Ethical Considerations

Participants were informed of the study’s purpose and assured full confidentiality. Participation was voluntary, with the right to withdraw at any point. No personally identifiable information was collected or recorded.

## Results

This chapter presents the findings derived from the study, which employed a mixed-methods approach. Quantitative data were collected via a survey administered to 172 postgraduate students and analyzed using descriptive statistics (frequencies and percentages). Concurrently, qualitative data were gathered through 25 semi-structured interviews and analyzed using thematic analysis to identify key themes. This chapter will first cover the quantitative demographic characteristics, followed by the survey results, and finally, the qualitative themes extracted from the interviews, to provide a comprehensive understanding of the topic.

### 1. Quantitative Results

The quantitative data collected from 172 postgraduate students across various Iraqi universities provides valuable insights into the challenges, practices, and perceptions surrounding the handling and exchange of biological samples. The findings are presented below according to key thematic areas of the study:

### Demographic Characteristics

To provide a comprehensive understanding of the sample, the demographic characteristics of the participants were analyzed. Figure 1 clearly illustrates the sample’s distribution in terms of gender and study level.

**Figure 1:**
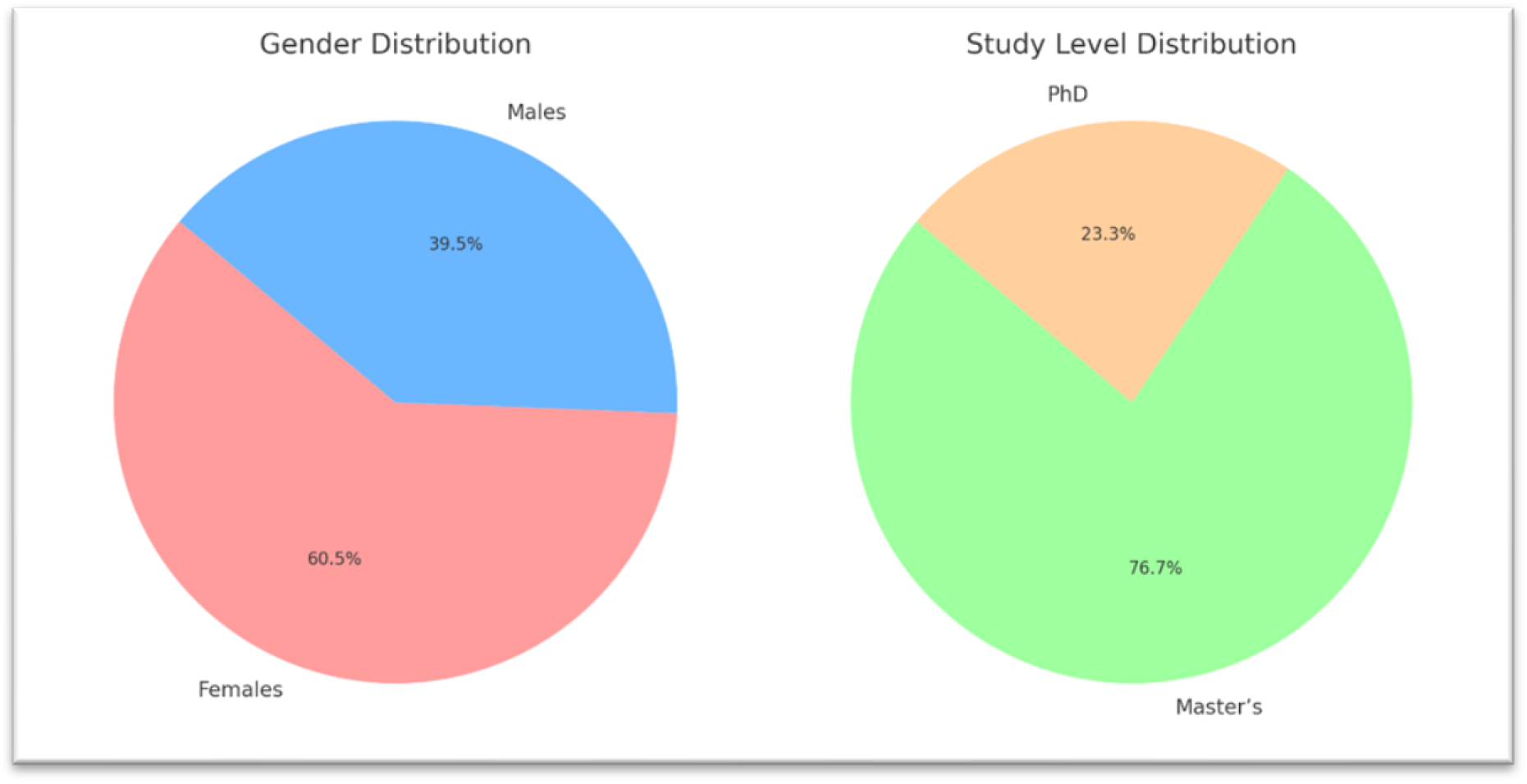
illustrates the sample’s distribution by gender and study level.

Regarding gender, females constituted the majority of participants, representing 60.5%, while males accounted for 39.5%. As for study level, Master’s students formed the largest proportion of the sample at 76.7%, with PhD students making up 23.3%.

#### 1. Challenges Related to Research and Resource Access

The results indicate that researchers encounter significant challenges in their research endeavors. As presented in Table 1, the majority of participants (71.5%) reported facing difficulty in obtaining biological samples to complete their research. Furthermore, 78.5% of respondents indicated challenges in completing their research due to time constraints or limited access to well-equipped university labs, with an additional 20.3% encountering these issues sometimes.

**Table 1:** Challenges Faced by Researchers in Sample Acquisition and Lab Access.

| Question | Answer Options | Count | Percentage (%) |
| --- | --- | --- | --- |
| 1. Have you ever faced difficulty obtaining biological samples to complete your research? | Yes | 123 | 71.5 |
|  | No | 5 | 2.9 |
|  | Sometimes | 44 | 25.6 |
| 2. Do you face difficulty completing your research due to time constraints or limited access to well-equipped university labs? | Yes | 135 | 78.5 |
|  | No | 2 | 1.2 |
|  | Sometimes | 35 | 20.3 |

#### 2. Sample Handling Practices and Reliance on External Labs

Findings revealed a significant reliance on external laboratories and sample exchange among colleagues (see Table 2). 65.1% of researchers reported using commercial labs or external offices for their experiments, while 80.2% had transferred samples from university labs to these external facilities to complete their research. The results also showed that 51.7% of participants had exchanged samples with colleagues to aid their research.

**Table 2:** Researchers’ Practices in Sample Usage, Transfer, and Exchange.

| Question | Answer Options | Count | Percentage (%) |
| --- | --- | --- | --- |
| 3. Do you use commercial labs or external offices to conduct your experiments or tests? | Yes | 112 | 65.1 |
|  | No | 12 | 7.0 |
|  | Sometimes | 48 | 27.9 |
| 4. Have you ever transferred samples from university labs to commercial labs or external offices to complete your research? | Yes | 138 | 80.2 |
|  | No | 1 | 0.6 |
|  | Sometimes | 33 | 19.2 |
| 5. Have you ever exchanged samples with colleagues to help them complete their research? | Yes | 89 | 51.7 |
|  | No | 50 | 29.1 |
|  | Sometimes | 33 | 19.2 |

#### 3. Awareness and Concerns Regarding Sample Integrity Risks

The survey demonstrates a high level of awareness regarding the risks associated with sample handling (see Table 3). An overwhelming majority of participants (91.3%) were aware that transferring samples between commercial labs or students might pose risks to sample integrity or result accuracy. Furthermore, 60.5% of researchers expressed concern about potential leakage or loss of samples during transport or exchange. Moreover, 82.0% of respondents categorized the exchange of samples between students and commercial labs as ‘high risk,’ while 60.5% believed that commercial labs might share their research samples with other students.

**Table 3:** Researchers’ Awareness and Concerns Regarding Sample Handling Risks.

| Question | Answer Options | Count | Percentage (%) |
| --- | --- | --- | --- |
| 6. Are you aware that transferring samples between commercial labs or students might pose risks to sample integrity or result accuracy? | Yes | 157 | 91.3 |
|  | No | 3 | 1.7 |
|  | Maybe | 12 | 7.0 |
| 7. Have you ever felt concerned about potential leakage or loss of samples during transport between labs or exchange among students? | Yes | 104 | 60.5 |
|  | No | 30 | 17.4 |
|  | Maybe | 38 | 22.1 |
| 8. What level of risk do you associate with exchanging samples between students and commercial labs? | High risk | 141 | 82.0 |
|  | Medium risk | 15 | 8.7 |
|  | Low risk | 15 | 8.7 |
|  | No risk | 1 | 0.6 |
| 9. Do you believe commercial labs may share your research samples with other students? | Yes | 104 | 60.5 |
|  | No | 12 | 7.0 |
|  | Maybe | 56 | 32.6 |

#### 4. Training, Protocols, and Support for Mandatory Training Programs

Regarding training in sample handling, findings revealed that 70.9% of participants had received practical training in biosafety within their universities (see Table 4). Of these, 55.2% received training during their undergraduate studies, 10.5% during postgraduate studies, and 29.1% through special courses. However, 41.3% of respondents reported a lack of written instructions or clear protocols for handling biological samples. A substantial 64.0% believed that some students begin working with samples without prior training. Concerning future support, the results indicated strong endorsement for mandatory training programs, with 69.8% of participants agreeing on the necessity of implementing such programs before allowing any student to handle biological samples.

**Table 4:** Researchers’ Perceptions and Practices Concerning Training and Protocols.

| Question | Answer Options | Count | Percentage (%) |
| --- | --- | --- | --- |
| 11. Have you ever received practical training within your university on how to handle biological samples as part of biosafety and biosecurity courses? | Yes | 122 | 70.9 |
|  | No | 46 | 26.7 |
|  | No, because my field is different | 4 | 2.3 |
| 12. If you received training in biosafety, where did it take place? | During undergraduate (BSc) | 95 | 55.2 |
|  | During postgraduate studies | 18 | 10.5 |
|  | Special courses | 50 | 29.1 |
|  | I didn't receive training because my field is different | 9 | 5.2 |
| 13. Were written instructions or clear protocols provided for handling biological samples? | Yes | 50 | 29.1 |
|  | No | 71 | 41.3 |
|  | Maybe | 51 | 29.7 |
| 14. Do you think some students start working with samples without prior training? | Yes | 110 | 64.0 |
|  | No | 13 | 7.6 |
|  | Maybe | 49 | 28.5 |
| 15. Would you support a mandatory training program before allowing any student to handle biological samples? | Yes | 120 | 69.8 |
|  | No | 15 | 8.7 |
|  | Maybe | 37 | 21.5 |

### Qualitative Results

This section presents the findings from the qualitative phase of the study, which involved 25 semi-structured interviews with postgraduate students. These students were drawn from diverse biological and medical science disciplines across various Iraqi universities. The interviews aimed to comprehensively explore the realities of biological sample handling, including motivations and challenges, awareness of associated risks, and the current state of biosafety training. The data were analyzed using Thematic Analysis to identify recurring patterns and insights from participants’ responses.

### Theme 1: Reasons Behind the Exchange of Samples Between Students and Commercial Laboratories

Analysis of participants’ responses revealed several consistent reasons driving the exchange of samples between students and commercial laboratories. These reasons primarily stemmed from systemic and resource-related limitations within the university environment: Time constraints and difficulties in completing research within the stipulated period were significant factors, articulated in 17 out of 25 interviews. Prohibitions on sample removal from hospitals, enforced by Ministry of Health regulations, were cited in 14 interviews, compelling students to seek external solutions.

Poor infrastructure in university laboratories and a critical shortage of essential materials emerged as a dominant concern, mentioned in 21 interviews. Reliance on commercial laboratories for quick results or the purchase of ready-made samples was also a common practice, noted in 19 interviews.

A typical sentiment encapsulating these challenges was expressed by a postgraduate student from the University of Baghdad:

> **“We don’t have the necessary materials, and the university labs are barely functioning for a few hours during official hours—if the equipment even works at all—so we have no choice but to go to commercial labs.”**

This quote underscores the direct link between inadequate university resources and the necessity of external lab reliance.

### Theme 2: Sample Exchange Practices Among Students

The interviews also unveiled a prevalent, albeit informal, practice of sample exchange among students. Some participants openly acknowledged sharing samples to help colleagues or to take advantage of previously examined specimens, especially in molecular research where time or resources may be scarce. As an example, one participant said:

> **“We don’t have time to start from scratch, so we take samples from colleagues who have already finished the practical part.”**

Others described this as a form of “informal exchange” primarily occurring within student groups, often driven by intense time pressure and the scarcity of available samples. Overall, 17 out of 25 participants admitted to personally engaging in sample exchange, while the remaining participants were aware of others doing so.

### Theme 3: Awareness of Biological Risks Associated with Sample Handling

Participants’ responses regarding awareness of biological risks varied, ranging from partial acknowledgment to a notable lack of concern. While 16 participants recognized the inherent biological risks, they often viewed sample exchange as an unavoidable reality within their research context. Some even expressed a belief that the risk was “exaggerated” if samples were handled with “care.” This perspective was highlighted by a student from the University of Karbala:

> **“I know it’s wrong, but I have no choice, and time is running out.”**

This statement reflects the tension between perceived risk and practical exigencies.

### Theme 4: Status of Training and Biosafety Awareness

A critical finding was the widespread admission by most students that they had not received proper biosafety training. Specifically, 15 interviews indicated a distinct lack of clear or structured training programs. When training did exist, it was often described as: Purely theoretical, lacking practical components. Outdated, with participants having received it during their undergraduate years. Non-specialized, failing to address specific risks pertinent to their current research. One participant’s statement clearly articulated this deficiency:

> **“The training was just a lecture in undergrad, with no practical component. We never learned how to handle dangerous samples properly.”**

### Theme 5: Students’ Suggestions and Recommendations

Participants offered several key recommendations aimed at improving sample management and biosafety practices within the academic research environment: Establishing official exemptions for postgraduate students to transfer samples under clearly defined protocols.

Improving university laboratories through better infrastructure, adequate supply of essential materials, and the implementation of an on-call lab system to extend operational hours.

Developing a national training program on biosafety and biosecurity to ensure standardized and comprehensive education. Providing students with legal and practical flexibility during the execution of their research, acknowledging the complexities of fieldwork and sample acquisition.

## Discussion of Findings

This mixed-methods study aimed to assess the risks associated with the unregulated handling of biological samples among postgraduate students in Iraq. The integrated quantitative and qualitative findings offer a comprehensive understanding of the field’s realities, illuminating existing challenges, prevalent practices, risk perceptions, and their subsequent implications for biosafety and biosecurity.

### Institutional Challenges as Drivers for Unregulated Practices

The quantitative results indicated that a substantial majority of participants (71.5% out of 172 students) faced difficulties in obtaining necessary biological samples, while 78.5% reported that time constraints and limited access to adequately equipped university laboratories hindered their research progress. These figures are deeply contextualized and elaborated by the qualitative analysis. Interviews revealed that “poor infrastructure in university labs and shortage of essential materials” was the most frequently cited reason (mentioned in 21 out of 25 interviews), directly compelling students to seek alternative solutions. A typical quote from a postgraduate student at the University of Baghdad encapsulates this reality: “We don’t have the necessary materials, and the university labs are barely functioning for a few hours during official hours—if the equipment even works at all—so we have no choice but to go to commercial labs.”

This resource deficit directly translates into reliance on “commercial laboratories and external offices to conduct experiments” (quantitatively acknowledged by 65.1% of students) and the transfer of samples to these external facilities (80.2% quantitatively). The absence of adequate infrastructure is not merely an academic impediment; it represents a critical biosafety vulnerability (Morse, 2025b; Moritz & Gillum, 2021). Once biological samples leave a controlled laboratory environment, they become susceptible to multiple risks, including contamination, loss, or unauthorized use (Petrillo, 2011). The lack of established protocols for transferring these samples between institutions, as evidenced by these practices, compromises the chain of custody, potentially leading to inaccurate research outcomes and increasing the likelihood of exposure to hazardous biological agents outside formal oversight (Kane et al., 2011), This unregulated movement of samples also raises biosecurity concerns, as it bypasses institutional controls designed to prevent the misappropriation or misuse of sensitive biological materials (Abad, 2012; Rainer & Cook, 2016).

### Peer-to-Peer Sample Exchange: Informal Collaboration with Inherent Risks

According to quantitative data, 51.7% of students acknowledged sharing biological samples with their peers, and another 19.2% did so on occasion. This suggests that informal collaboration is a coping mechanism. This behavior is supported by qualitative data, as 17 out of 25 participants personally admitted to exchanging samples, while the remaining participants said they knew others who did so. Statements like “We take samples from colleagues who already completed the practical part, because we don’t have time to start from scratch” demonstrate how students are compelled to find these easy fixes due to time constraints and a lack of available samples.

While seemingly innocuous from the students’ perspective, this practice poses significant biosafety and biosecurity risks. Informal exchange means a complete absence of traceability records, making it difficult to pinpoint the sample’s origin in cases of contamination or misuse (Smith & Sandbrink, 2022; Ganesh et al., 2023). It also elevates the risk of pathogen transmission among individuals if stringent safety procedures (e.g., proper personal protective equipment use, safe waste disposal) are not followed (Golovko & Napnenko, 2024; Jalali et al., 2024). From a biosecurity standpoint, this signifies a loss of control over potential biological agents, which could lead to undeclared laboratory incidents or even the unintentional release of pathogens into the community, thereby threatening public health (Blacksell et al., 2023a; Golovko & Napnenko, 2024).

### Risk Awareness Versus Reality Pressures: A Biosafety Dilemma

The discrepancy between high-risk awareness and ongoing participation in risky behaviors is a startling paradox in this study. 91.3% of students acknowledged, quantitatively, that sample transfer would jeopardize sample integrity and result accuracy, while 82% said sample exchange involved “high risk.” However, 60.5% of respondents still engaged in these procedures despite expressing concern about possible sample loss or leakage.

Qualitative findings elucidate this cognitive and behavioral dissonance. While 16 participants acknowledged the biological risks, they perceived them as an “unavoidable reality.” A student from the University of Karbala vividly expressed this dilemma: “I know it’s wrong, but I have no choice, and time is running out.” This highlights that awareness alone is insufficient to alter behaviors if viable alternatives are not available. In terms of biosafety, this means students’ knowledge of risks does not translate into safe practices due to academic and institutional pressures, creating a dangerous gap between awareness and the actual implementation of biosafety measures (Thi Ngoc Ha Bui et al., 2024). This significantly increases the likelihood of serious biological incidents, given inadequate training and protocols (Blacksell et al., 2023b; Padde et al., 2022).

### Training and Protocol Gaps: A Fragile Foundation for Biosecurity

Although 70.9% of students reported receiving some form of practical instruction, qualitative results revealed the insufficient nature of this training. Fifteen interviews indicated a lack of clear training programs, with existing training described as “purely theoretical, outdated (from undergraduate years), or non-specialized.” One participant elaborated: “The training was just a lecture in undergrad, with no practical component. We never learned how to handle dangerous samples properly.”

This issue is compounded by the scarcity of clear written protocols for biological sample handling, confirmed by only 29.1% of students. These gaps in training and protocols pose a substantial biosafety and biosecurity risk (Alrasheed et al., 2024b; Laboratory Specialist, Ministry of Health, Saudi Aribia & Alansari, 2023). The absence of specialized practical training means students may lack fundamental skills for safe sample handling, increasing the risk of exposure to pathogens, cross-contamination, and infection spread (Azaev et al., 2023; Sveinbjornsson & Gizurarson, 2022b). Furthermore, the lack of standardized protocols fosters disorganization, makes sample tracking difficult, and creates an environment conducive to biological incidents and the potential misuse of biological materials (Samuel, 2018; Verma & Yadav, 2023). The strong support (69.8%) for implementing a mandatory training program underscores students’ recognition of this critical gap and the urgent need for institutional intervention.

### Study Limitations

#### Uneven Representation of Universities

Due to the online distribution of the survey, participation varied significantly among different universities. This imbalance may have influenced the comprehensiveness of the overall picture.

#### Descriptive Statistics Only

The study relied solely on frequencies and percentages for data analysis. Inferential statistics were not applied due to the uneven institutional representation, which limited deeper comparisons between variables.

#### Interview Documentation Method

For the qualitative part, interviews were not audio-recorded. Instead, the researcher took notes during conversations to ensure participant comfort and privacy. While practical, this approach may have led to the loss of some nuanced details.

#### Sensitivity of the Topic

Given the nature of the subject—handling biological samples outside formal protocols—some participants may have been hesitant to speak openly, which might have affected the richness or honesty of some responses.

## Data Availability

All data produced in the present study are available upon reasonable request to the authors

